# Points-Based Models for Predicting Severe Disease related to Covid-19 with the SARS-CoV-2 Omicron Variant

**DOI:** 10.1101/2022.06.27.22276907

**Authors:** Yatir Ben Shlomo, Jacob G. Waxman, Galit Shaham, Doron Netzer, Ben Reis, Ran Balicer, Noa Dagan

**Affiliations:** Clalit Research Institute, Innovation Division, Clalit Health Services, Tel Aviv, Israel; Community Medical Services Division, Clalit Health Services, Arlozorov 101, Tel Aviv, Israel; Predictive Medicine Group, Computational Health Informatics Program, Boston Children’s Hospital, Boston, MA, USA; Harvard Medical School, Boston, MA, USA; School of Public Health, Faculty of Health Sciences, Ben Gurion University of the Negev, Be’er Sheva, Israel; Software and Information Systems Engineering, Ben Gurion University, Be’er Sheva, Israel; Department of Biomedical Informatics, Harvard Medical School, Boston, MA, USA

## Abstract

**Introduction:** Throughout the SARS-CoV-2 pandemic, resources for various aspects of patient care have been limited, necessitating risk-stratification. The need for good risk-stratification tools has been enhanced by the availability of new Covid-19 therapeutics that are effective at preventing severe disease among high-risk patients if given promptly following SARS-CoV-2 infection. We describe the development of two points-based models for predicting the risk of deterioration to severe disease from an Omicron-variant SARS-CoV-2 infection.

**Methods:** We developed two logistic regression-based models for predicting the risk of severe Covid-19 within a 21-days follow-up period among Clalit Health Services members aged 18 and older, with confirmed SARS-CoV-2 infection from December 25, 2021 to March 16, 2022. In the first model, aimed for the use of healthcare providers, the model coefficients were linearly transformed into integer risk points. In the second model, a simplified version designed for self-assessment by the general public, the risk points were further scaled down to smaller numbers with less variability across risk factors.

**Results:** 613,513 individuals met the inclusion criteria, of which 1,763 (0.287%) developed the outcome. The AUROC estimates for both models were 0.95, although the ‘full’ model demonstrated more granular risk-stratification capabilities (77 vs. 27 potential thresholds on the test set). Both models proved effective in identifying small subsets of the population enriched with individuals who ended up deteriorating. For example, prioritizing the top 1%, 5% or 10% individuals in the population for interventions with the full model results in coverage of 36%, 68% or 83% (respectively) of the individuals that actually end up deteriorating. Risk point count increased with age, number of chronic conditions and previous hospitalizations, and decreased with recent vaccination and infection.

**Discussion:** The models presented, one more expressive and one more accessible, are transparent and explainable models applicable to the general population that can be used in the prioritization of Covid-19-related resources, including therapeutics.

## Introduction

Despite widespread and successful vaccination campaigns, novel variants and waning of vaccine-induced immunity are perpetuating the SARS-CoV-2 pandemic. (1) Novel Covid-19 therapeutics are a limited resource due to their cost and the challenging logistics of administering them promptly following infection diagnosis, delivering the medication directly to the homes of patients with confirmed infections. As a result, there is a need for risk prioritization that will facilitate identification of the patients most at need for these treatments on a daily basis.

The development of prediction models requires balancing the complexity or expressivity of the model, on the one hand, with its explainability or transparency on the other hand. Higher expressivity is generally associated with higher performance and greater variability of risk estimations. This usually translates to a model with high granularity of risk cut-offs that can be used to classify the population into increased or regular risk groups. At the extreme end of the expressivity spectrum are machine-learning models that can be based on an extremely large number of variables.

However, these models are “black-boxes”, and this poses a challenge when prioritizing sensitive resources that requires models to be explainable and transparent (“white-boxes” that can be easily understood). Transparency was a central theme in the early discussions surrounding prioritization of Covid-19 vaccines (2). Due to the expressivity-explainability tradeoff, developers must recognize the importance of these properties within the context of the model’s designated use.

Almost two years into the Covid-19 pandemic, Clalit Health Services (CHS), a healthcare organization responsible for the care of over half of the Israeli population, has deployed and used Covid-19 prediction models in both extremes of the of the expressivity-explainability continuum(3,4). For the purpose of prioritization of sensitive, limited resources such as Covid-19 therapeutics, that are available in various and changing amounts, it was recognized that there is a need to develop models that will be explainable and transparent, while providing good performance with enough granularity to enable allocation of resources dynamically across various risk cut-offs. A points-based scoring system that augments and simplifies a more expressive raw model has the benefit of being both adequately expressive, while maintaining transparency and explainability, representing a middle point in the expressivity-explainability continuum.

This paper describes the development and performance of two points-based logistic regression-derived models for the allocation of Covid-19 therapeutics on a daily basis. Although both models are transparent and provide good performance, they still differ in the level of tradeoff between explainability and granularity. The ‘full’ model, targeted for use by healthcare providers, has the highest possible fidelity with the underlying raw logistic regression model, maximizing its risk-stratification capabilities. The ‘simplified’ model, which aims to enable self-assessment by the general public – an important tool for policymaker-public communication, maximizes ease of calculation at the expense of expressivity and granularity.

## Methods

### Data source

The Models were trained with data from CHS, the largest integrated payer-provider healthcare organization in Israel. The CHS data repositories include over 4.8 million members, accounting for approximately 53% of the Israeli population. These data repositories contains both inpatient and outpatient data, which enables CHS to ascertain a full medical history of its members. COVID-19 related data such as diagnostic test results, hospitalizations and disease severity, are collected centrally by the Israeli Ministry of Health (MOH) and sent daily to the respective health care provider.

### Study population

The derivation population used for developing the models included all individuals aged 18 and above, who were identified as covid-19 positives (excluding borderline cases) and did not receive any medication for preventing severe COVID-19 outcome, during December 25, 2021 through March 16, 2022. This period marks the Omicron surge in Israel.

### Outcomes and model variables

The outcome of interest was severe Covid-19, defined according to the Israeli ministry of health criteria (including events of Covid-19-related deaths). The follow-up period for determining the outcome was 21 days from the date of confirmed infection.

The outcome was modeled using a list of known risk factors and prognostic variables. The following variables were introduced into the model: demographic factors; known or possible risk factors for severe Covid-19 as defined by the Centers of Diseases Control and Prevention(5); amount of hospitalizations in recent years as a marker for burden and stability of chronic conditions; and immunization status (vaccination and previous infection). Due to evidence of waning immunity over time(6), the effect of Covid-19 vaccination was included as a variable quantifying the time that has elapsed since the individual’s latest vaccine dose (second, third or fourth).

### Statistical Analysis

The outcome was modeled using logistic regression. The final decision of which variables to include in the model was based on statistical significance of effects; preventing multi-collinearity; ease of determining the value of each variable for self-assessment of risk scores by CHS members; and fairness considerations.

Multi-collinearity status was evaluated by calculating the Variable Inflation Factor (VIF) for each covariate in the model. The model was fitted on a random subset of 80% of the dataset (the train set), while the remaining 20% was kept for model evaluation (the test set).

#### Full Points Model

The raw model coefficients were transformed to risk points (integer values) via multiplication by a constant factor and rounding the result. This constant factor was chosen such that the smallest statistically significant raw model coefficient, in absolute value, was transformed to a single risk point. The final risk score is calculated by summation of risk points of all the model variables. Since certain conditions, such as recent vaccination, decrease the predicted risk of the outcome, the resulting model has the potential to assign a negative risk score for the young, healthy, vaccinated individuals. In order to prevent negative scores, the associated risk points of all levels of the age group variable (including the reference level) was increased accordingly by a constant number, such that the minimal possible risk score generated by the model is zero points.

#### Simplified Points Model

In order to enable easy and accurate self-assessment of risk, an additional model was derived by including an additional processing step, in which raw model coefficients were divided by 3 prior to their being rounded to integer values. Resulting values (after division by 3) were capped to a minimal value of 1 (or -1 for negative coefficients) in order to prevent a variable from being associated with 0 risk points. Accordingly, risk points 0.5 - 4.49 were simplified to 1, risk points 4.5 – 7.49 were simplified to 2 and so forth, with similar transformation occurring for negative risk points.

#### Missing values

Other than obesity status, no variables had missing values. For individuals with missing BMI (required to define the obesity status), imputation was performed with obesity status set to 0.

#### Evaluation of Model Performance

Both the full and simplified points models were evaluated on the test dataset. The models were evaluated for area under the receiver operator curve (AUROC). In addition, for each potential threshold (i.e. each unique risk score the model generated) the following metrics were calculated: sensitivity, positive predictive value (PPV) and lift.

#### Sensitivity Analysis

The exclusion of patients who received Covid-19 therapy might introduce bias in estimating the risk for the highest risk patients (who are more likely to receive this therapy). To examine the effect on the resulting model, a separate analysis included these patients as a sensitivity analysis.

## Results

A total of 613,513 individuals met the inclusion criteria: aged 18 and above, a confirmed SARS-CoV-2 infection between December 25, 2021 and March 16, 2022, and continuous CHS membership (Supplementary Figure 1). A total of 1.1% had missing BMI value that was imputed as no obesity.

1,763 (0.287%) individuals developed the outcome during the follow-up period. The dataset used for training had 490,810 observations, 122,703 observations were set aside for the purpose of model evaluation. Baseline characteristics of the dataset with stratification by outcome are described in Table 1.

**Table 1:** Study population, stratified by outcome

| Variable | Overall, N = 613,513<sup>1</sup> | Non-Severe Outcome,<br>N = 611,750 (99.7%)<sup>1</sup> | Severe Outcome,<br>N = 1,763 (0.3%)<sup>1</sup> |
| --- | --- | --- | --- |
| **Age group** |  |  |  |
| 18 - 29 | 115,826 (19%) | 115,812 (19%) | 14 (0.8%) |
| 30 - 39 | 167,854 (27%) | 167,829 (27%) | 25 (1.4%) |
| 40 - 49 | 142,262 (23%) | 142,203 (23%) | 59 (3.3%) |
| 50 - 59 | 78,133 (13%) | 78,001 (13%) | 132 (7.5%) |
| 60 - 69 | 61,225 (10.0%) | 60,946 (10.0%) | 279 (16%) |
| 70 - 79 | 32,426 (5.3%) | 31,954 (5.2%) | 472 (27%) |
| 80+ | 15,787 (2.6%) | 15,005 (2.5%) | 782 (44%) |
| **Sex** |  |  |  |
| Female | 362,845 (59%) | 362,041 (59%) | 804 (46%) |
| Male | 250,668 (41%) | 249,709 (41%) | 959 (54%) |
| **Obesity** | 124,482 (20%) | 123,852 (20%) | 630 (36%) |
| **Active cancer** | 16,117 (2.6%) | 15,799 (2.6%) | 318 (18%) |
| **Chronic Kidney Disease** | 11,717 (1.9%) | 11,451 (1.9%) | 266 (15%) |
| **Diabetes Mellitus** | 19,721 (3.2%) | 19,450 (3.2%) | 271 (15%) |
| **Heart disease** | 16,315 (2.7%) | 15,844 (2.6%) | 471 (27%) |
| **Neurological disease** | 9,167 (1.5%) | 8,928 (1.5%) | 239 (14%) |
| **Liver disease** | 3,190 (0.5%) | 3,130 (0.5%) | 60 (3.4%) |
| **Chronic Obstructive Pulmonary Disease** | 2,098 (0.3%) | 2,027 (0.3%) | 71 (4.0%) |
| **Immunosuppression** | 21,105 (3.4%) | 20,775 (3.4%) | 330 (19%) |
| **Number of hospital admissions**<br>**in previous 3 years** |  |  |  |
| 0 | 511,959 (83%) | 511,384 (84%) | 575 (33%) |
| 1 - 2 | 87,057 (14%) | 86,469 (14%) | 588 (33%) |
| 3 - 5 | 11,906 (1.9%) | 11,528 (1.9%) | 378 (21%) |
| 6+ | 2,591 (0.4%) | 2,369 (0.4%) | 222 (13%) |
| **Months from last vaccine dose**<br>**(2<sup>nd</sup>, 3<sup>rd</sup> or 4<sup>th</sup>)** |  |  |  |
| unvaccinated | 102,763 (17%) | 102,073 (17%) | 690 (39%) |
| 1 - 6 | 414,174 (68%) | 413,383 (68%) | 791 (45%) |
| 7 - 10 | 34,982 (5.7%) | 34,871 (5.7%) | 111 (6.3%) |
| 11+ | 61,594 (10%) | 61,423 (10%) | 171 (9.7%) |
| **Previous infection** | 55,093 (9.0%) | 54,949 (9.0%) | 144 (8.2%) |

<sup>1</sup>n (%)

In preliminary analysis the following variables were considered, but excluded from the models due to small or non-significant effect: pregnancy, hypertension, smoking status, and asthma. The binary variable ‘sex’ was removed due to fairness considerations. The categorical variable ‘number of vaccine doses’ was removed to prevent collinearity with the variable of ‘months from last Covid-19 vaccine dose’. The final model variables include: age group, obesity, active cancer, chronic kidney disease (CKD), diabetes of any type, heart disease)including cerebrovascular disease), neurological disease, liver disease, chronic obstructive pulmonary disease, immunosuppression, number of hospitalizations in the 3 years prior to the index date (as a categorical variable: 0, 1-2, 3-5, 6 or more), previous Covid-19 infection (as a binary variable), months from last Covid-19 vaccination dose (provided that at least 2 doses were administered, as a categorical variable: unvaccinated, 1-6, 7-10, 11 or more). The highest VIF value for the final model variables was 2.2, indicating that there was no evidence of any further multi-collinearity.

The logistic regression model coefficients are specified in supplementary Table S1. Age is the single most important predictive risk factor for developing a severe outcome. Indeed, a clear, monotonically increasing, association between age and the outcome is evident. The most important prognostic variables following age were: number of hospitalizations and the time from last vaccination dose. Both variables show clear monotonic trends with the outcome.

The final full and simplified point-based models are described in Table 2. Test set performance metrics are presented on Tables 3 and 4, showing the predicted risk associated with each risk score, as well as the percent of population above each possible risk score serving as a threshold, for the full and simplified models respectively. The full model demonstrated more granular risk-stratification capabilities, offering 77 potential thresholds compared to 27 potential thresholds of the simplified model. For dynamic resource allocation that includes prioritizing shares of up to 20% of the population, the full models offers risk-thresholds with maximal increase of 2% between thresholds. The simplified model offers numerous thresholds as well, but the maximal increase in this range is 4%, offering less granularity.

**Table 2:** Full and simplified point models – allocation of points per risk factor

| **Variable** | **Full Points Model**<br>**points allocation** | **Simplified Points Model**<br>**points allocation** |
| --- | --- | --- |
| **Age group** |  |  |
| 18-29 | 21 | 7 |
| 30-39 | 25 | 8 |
| 40-49 | 31 | 10 |
| 50-59 | 39 | 13 |
| 60-69 | 45 | 15 |
| 70-79 | 51 | 1 |
| 80+ | 57 | 19 |
| **Obesity** | 2 | 1 |
| **Active cancer** | 3 | 1 |
| **Chronic Kidney Disease** | 3 | 1 |
| **Diabetes Mellitus** | 1 | 1 |
| **Heart disease** | 1 | 1 |
| **Neurological disease** | 2 | 1 |
| **Liver disease** | 3 | 1 |
| **Chronic Obstruction**<br>**Pulmonary Disease** | 2 | 1 |
| **immunosuppression** | 5 | 2 |
| **Number of hospital**<br>**admissions in previous 3 years** |  |  |
| 1 - 2 | 5 | 2 |
| 3 - 5 | 10 | 3 |
| 6+ | 13 | 4 |
| **Months from last vaccine dose**<br>**(2nd, 3rd or 4th)** |  |  |
| 1 - 6 | -14 | -5 |
| 7 - 10 | -10 | -3 |
| 11+ | -7 | -2 |
| **Previous infection** | -7 | -2 |

**Table 3:** Test set performance metrics of the full model for all possible risk scores (thresholds)

| Points | Minimum predicted risk* | Maximum predicted risk* | % population above threshold | Sensitivity | PPV | Lift |
| --- | --- | --- | --- | --- | --- | --- |
| 0 | 0.001% | 0.001% | 100% | 1 | 0.003 | 1.001 |
| 2 | 0.001% | 0.001% | 99.85% | 1 | 0.003 | 1.002 |
| 3 | 0.002% | 0.002% | 99.84% | 1 | 0.003 | 1.002 |
| 4 | 0.002% | 0.002% | 99.84% | 1 | 0.003 | 1.004 |
| 5 | 0.002% | 0.002% | 99.63% | 0.997 | 0.003 | 1.001 |
| 6 | 0.002% | 0.002% | 99.61% | 0.997 | 0.003 | 1.001 |
| 7 | 0.003% | 0.003% | 99.58% | 0.997 | 0.003 | 1.093 |
| 8 | 0.003% | 0.003% | 91.24% | 0.997 | 0.003 | 1.093 |
| 9 | 0.004% | 0.004% | 91.20% | 0.997 | 0.003 | 1.104 |
| 10 | 0.004% | 0.005% | 90.29% | 0.997 | 0.003 | 1.107 |
| 11 | 0.005% | 0.006% | 90.04% | 0.997 | 0.004 | 1.302 |
| 12 | 0.006% | 0.007% | 76.60% | 0.994 | 0.004 | 1.315 |
| 13 | 0.007% | 0.008% | 75.60% | 0.994 | 0.004 | 1.354 |
| 14 | 0.007% | 0.009% | 73.42% | 0.994 | 0.004 | 1.424 |
| 15 | 0.009% | 0.011% | 69.82% | 0.994 | 0.004 | 1.445 |
| 16 | 0.010% | 0.012% | 68.81% | 0.992 | 0.004 | 1.489 |
| 17 | 0.012% | 0.015% | 66.62% | 0.989 | 0.005 | 1.795 |
| 18 | 0.014% | 0.018% | 55.07% | 0.989 | 0.006 | 1.949 |
| 19 | 0.016% | 0.021% | 50.74% | 0.989 | 0.006 | 2.075 |
| 20 | 0.019% | 0.025% | 47.66% | 0.989 | 0.006 | 2.13 |
| 21 | 0.022% | 0.031% | 46.42% | 0.986 | 0.007 | 2.271 |
| 22 | 0.025% | 0.033% | 43.42% | 0.983 | 0.007 | 2.332 |
| 23 | 0.030% | 0.040% | 42.16% | 0.975 | 0.007 | 2.37 |
| 24 | 0.036% | 0.046% | 41.14% | 0.975 | 0.007 | 2.542 |
| 25 | 0.041% | 0.057% | 38.35% | 0.972 | 0.009 | 3.175 |
| 26 | 0.047% | 0.068% | 30.62% | 0.969 | 0.01 | 3.297 |
| 27 | 0.056% | 0.077% | 29.40% | 0.969 | 0.01 | 3.572 |
| 28 | 0.066% | 0.094% | 27.14% | 0.966 | 0.011 | 3.636 |
| 29 | 0.075% | 0.099% | 26.58% | 0.964 | 0.011 | 3.745 |
| 30 | 0.089% | 0.124% | 25.73% | 0.953 | 0.011 | 3.878 |
| 31 | 0.107% | 0.148% | 24.56% | 0.933 | 0.014 | 4.837 |
| 32 | 0.119% | 0.169% | 19.29% | 0.933 | 0.015 | 5.26 |
| 33 | 0.146% | 0.197% | 17.74% | 0.922 | 0.017 | 5.853 |
| 34 | 0.174% | 0.234% | 15.75% | 0.916 | 0.018 | 6.177 |
| 35 | 0.195% | 0.280% | 14.83% | 0.908 | 0.019 | 6.425 |
| 36 | 0.229% | 0.325% | 14.13% | 0.897 | 0.02 | 6.826 |
| 37 | 0.269% | 0.372% | 13.14% | 0.883 | 0.023 | 7.989 |
| 38 | 0.315% | 0.444% | 11.05% | 0.869 | 0.025 | 8.697 |
| 39 | 0.374% | 0.558% | 9.99% | 0.838 | 0.029 | 9.961 |
| 40 | 0.421% | 0.598% | 8.41% | 0.824 | 0.031 | 10.683 |
| 41 | 0.515% | 0.711% | 7.71% | 0.813 | 0.033 | 11.462 |
| 42 | 0.589% | 0.817% | 7.09% | 0.796 | 0.036 | 12.46 |
| 43 | 0.708% | 0.967% | 6.39% | 0.76 | 0.041 | 13.896 |
| 44 | 0.782% | 1.182% | 5.47% | 0.726 | 0.044 | 15.091 |
| 45 | 0.980% | 1.311% | 4.81% | 0.682 | 0.051 | 17.518 |
| 46 | 1.107% | 1.618% | 3.89% | 0.665 | 0.056 | 19.086 |
| 47 | 1.295% | 1.902% | 3.48% | 0.637 | 0.06 | 20.712 |
| 48 | 1.594% | 2.223% | 3.08% | 0.595 | 0.067 | 22.793 |
| 49 | 1.894% | 2.459% | 2.61% | 0.564 | 0.071 | 24.293 |
| 50 | 2.148% | 2.930% | 2.32% | 0.522 | 0.078 | 26.874 |
| 51 | 2.509% | 3.469% | 1.94% | 0.483 | 0.089 | 30.361 |
| 52 | 2.944% | 3.971% | 1.59% | 0.447 | 0.095 | 32.488 |
| 53 | 3.433% | 4.697% | 1.38% | 0.413 | 0.106 | 36.389 |
| 54 | 4.004% | 5.381% | 1.14% | 0.383 | 0.112 | 38.552 |
| 55 | 4.625% | 6.662% | 0.99% | 0.36 | 0.119 | 40.863 |
| 56 | 5.446% | 7.634% | 0.88% | 0.327 | 0.131 | 44.957 |
| 57 | 6.183% | 8.266% | 0.73% | 0.257 | 0.129 | 44.35 |
| 58 | 7.270% | 9.480% | 0.58% | 0.226 | 0.134 | 45.888 |
| 59 | 8.209% | 11.236% | 0.49% | 0.19 | 0.141 | 48.254 |
| 60 | 9.625% | 13.576% | 0.39% | 0.168 | 0.147 | 50.281 |
| 61 | 10.876% | 15.017% | 0.33% | 0.154 | 0.154 | 52.804 |
| 62 | 12.902% | 16.372% | 0.29% | 0.12 | 0.154 | 52.825 |
| 63 | 15.001% | 19.004% | 0.23% | 0.109 | 0.169 | 57.866 |
| 64 | 16.947% | 21.838% | 0.19% | 0.098 | 0.192 | 65.913 |
| 65 | 19.767% | 24.655% | 0.15% | 0.075 | 0.188 | 64.265 |
| 66 | 22.422% | 27.477% | 0.12% | 0.07 | 0.202 | 69.102 |
| 67 | 24.401% | 30.861% | 0.10% | 0.059 | 0.204 | 69.88 |
| 68 | 29.189% | 33.746% | 0.08% | 0.056 | 0.25 | 85.686 |
| 69 | 30.994% | 37.237% | 0.07% | 0.05 | 0.281 | 96.397 |
| 70 | 35.863% | 41.368% | 0.05% | 0.042 | 0.312 | 107.108 |
| 71 | 38.518% | 44.568% | 0.04% | 0.028 | 0.294 | 100.808 |
| 72 | 42.441% | 48.521% | 0.03% | 0.011 | 0.174 | 59.608 |
| 73 | 49.729% | 51.904% | 0.02% | 0.011 | 0.2 | 68.549 |
| 74 | 51.874% | 56.450% | 0.02% | 0.011 | 0.25 | 85.686 |
| 75 | 55.051% | 59.445% | 0.01% | 0.003 | 0.125 | 42.843 |
| 77 | 64.523% | 65.933% | 0.01% | 0.003 | 0.25 | 85.686 |

\* Minimum and maximum predicted risks are obtained from the underlying logistic regression model and represent the range of risk prediction for all observations in the test set that were assigned the corresponding risk score. Taken together they allow for mapping a given risk score to an absolute risk estimation.

**Table 4:** Test set performance metrics of the simplified model for all possible risk scores (thresholds)

| Points | Minimum predicted risk | Maximum predicted risk | % population above threshold | Sensitivity | PPV | Lift |
| --- | --- | --- | --- | --- | --- | --- |
| 0 | 0.001% | 0.001% | 100% | 1 | 0.003 | 1.001 |
| 1 | 0.001% | 0.002% | 99.85% | 1 | 0.003 | 1.003 |
| 2 | 0.002% | 0.003% | 99.66% | 0.997 | 0.003 | 1.092 |
| 3 | 0.002% | 0.005% | 91.35% | 0.997 | 0.004 | 1.283 |
| 4 | 0.004% | 0.009% | 77.70% | 0.994 | 0.004 | 1.354 |
| 5 | 0.005% | 0.014% | 73.42% | 0.989 | 0.005 | 1.763 |
| 6 | 0.008% | 0.024% | 56.08% | 0.989 | 0.006 | 2.063 |
| 7 | 0.009% | 0.041% | 47.94% | 0.983 | 0.007 | 2.323 |
| 8 | 0.017% | 0.065% | 42.33% | 0.972 | 0.009 | 3.141 |
| 9 | 0.028% | 0.109% | 30.95% | 0.966 | 0.01 | 3.581 |
| 10 | 0.050% | 0.183% | 26.99% | 0.941 | 0.014 | 4.757 |
| 11 | 0.068% | 0.241% | 19.79% | 0.922 | 0.017 | 5.863 |
| 12 | 0.096% | 0.416% | 15.72% | 0.888 | 0.022 | 7.562 |
| 13 | 0.157% | 0.700% | 11.75% | 0.838 | 0.028 | 9.744 |
| 14 | 0.269% | 1.108% | 8.60% | 0.777 | 0.037 | 12.686 |
| 15 | 0.368% | 1.621% | 6.12% | 0.679 | 0.048 | 16.363 |
| 16 | 0.600% | 2.632% | 4.15% | 0.617 | 0.064 | 21.917 |
| 17 | 0.823% | 4.283% | 2.82% | 0.508 | 0.083 | 28.562 |
| 18 | 1.246% | 6.978% | 1.78% | 0.394 | 0.104 | 35.509 |
| 19 | 2.295% | 11.236% | 1.11% | 0.285 | 0.122 | 41.669 |
| 20 | 3.694% | 16.372% | 0.68% | 0.184 | 0.134 | 45.978 |
| 21 | 5.770% | 21.556% | 0.40% | 0.126 | 0.153 | 52.461 |
| 22 | 8.809% | 28.086% | 0.24% | 0.081 | 0.176 | 60.24 |
| 23 | 15.490% | 40.305% | 0.13% | 0.059 | 0.236 | 80.873 |
| 24 | 22.176% | 45.867% | 0.07% | 0.034 | 0.267 | 91.399 |
| 25 | 29.219% | 59.445% | 0.04% | 0.017 | 0.3 | 102.824 |
| 26 | 42.441% | 64.648% | 0.02% | 0.003 | 0.167 | 57.124 |
| 27 | 64.523% | 73.963% | 0.01% | 0.003 | 0.5 | 171.373 |

The test-set AUROC of both the full and simplified models was 0.95. Tables 3 and 4 also present the test set performance metrics per each potential model threshold. Both models proved effective in identifying small subsets of the population that held high proportion of those who ended up deteriorating – for example, the full model offers prioritizing 1%, 5% or 10% of the population for interventions, capturing 36%, 68% or 83% of the high-risk patients.

### Sensitivity Analysis

To verify that excluding patients who received Covid-19 therapy did not introduce significant bias in the risk estimated by the models, we compared the resulting models when these individuals were included in the analysis to the models in the main analysis in which these individuals were excluded. The models are summarized in supplementary Table S2.

The differences are small and mostly amount to a single risk point difference in some of the variables. Discrepancies larger than a single risk points occurred only once, in the risk points for age group 30-39, where only 25 events are observed and the estimate is not statistically significant. The changes in test-set AUROC for the resulting models is 0.002 (0.948 vs 0.950).

## Discussion

We developed two related point-based models for adults, aged 18 years old and above, that accurately predict the risk of deterioration from infection with the Omicron variant of SARS-CoV-2 to severe Covid-19. Although both models are transparent and offer good granularity, they still vary in their level of simplicity, with the more complex model suitable for use by healthcare providers and the simpler one targeted for use by the general public. Whilst the more complex model is more expressive (as would be expected) and allows for more granular risk stratification, both models show very high levels of discrimination (with AUROC = 0.95 for both models).

Prioritization has played a critical role in this pandemic, and over the course of it CHS employed risk-stratification tools in numerous settings. For example, clinically-high-risk individuals, as defined by a predictive model, were targeted for proactive outreach by family care practitioners and nurses, with the aim of helping them minimize their risk. They did this by making these individuals aware of their risk and offering services such as home delivery of their regular medications to help them reduce their exposure risk(4). In addition, the predictive modes were used to prioritize patients for inpatient care once they were infected (in situations where it would have been clinically justifiable for them to be managed in the community). More recently, there was a need for prioritization of eligible candidates for novel Covid-19 therapeutics. Existing tools were either not transparent enough or did not offer enough granularity of thresholds(3,4)

A new targeted approach for optimizing the daily allocation of Covid-19 therapeutics was needed. Therefore, we leveraged CHS’s rich EMR to develop up-to-date and accurate models that take into account both vaccination status and known risk factors, all within the current variant ‘landscape’ predominated by Omicron. We put an emphasis on models that would be both transparent and suitable expressive. Most existing models for evaluating risk of severe Covid-19 were developed and published in the beginning of the pandemic and are not suitable for the prioritization of Covid-19 therapeutics(7) – they are either not transparent in a manner that allows for manual risk evaluating, do not take into account features such as Covid-19 vaccines or prior SARS-CoV-2 infection, or are based on a population of patients that were admitted to a hospital rather than the general population of infected individuals.

Our study is subject to a number of limitations. First, our analysis excludes individuals that received a Covid-19 therapy since high-risk individuals successfully treated with these therapeutics might be wrongly classified by the model as low risk and not prioritized correctly. This may bias our predictions towards underestimation since the highest risk patients were more likely to receive these preventative treatments. However, due to the limited supply, challenging logistics of administering these drugs, and varying adherence rates, the study cohort still held substantial representation of high-risk patients. In a sensitivity analysis, which included these treated subjects, the results were not significantly different in terms of the final point assignment and performance metrics (Supplementary Table S2).

Second, the model was trained on data acquired during the period in which the Omicron variant was predominant in Israel – a variant that has been shown to result in lower rates of severe outcomes relative to previous variants. Hence, the model might not be generalizable to novel future variants. Third, rare conditions associated with a high risk of deterioration to severe Covid-19 may be missed by the model due to lack of the required sample size for being allotted a point. This concern is applicable to all risk prediction models or risk factor lists. This concern could be mitigated somewhat by the inclusion of risk points for all-cause hospitalization, as well as enabling diversion of resources to individuals classified as low risk at the treating physician’s discretion.

In conclusion, we describe the development of two point-based models assessing risk of deterioration to severe Covid-19 or Covid-19-related death in individuals infected with the Omicron variant. These models are highly transparent, while maintaining good performance and sufficient granularity, and can aid in the daily prioritization of Covid-19.

## Supporting information

TRIPOD checklist

## Data Availability

Owing to data privacy regulations, the raw data for this study cannot be shared.

