## Supplemental data for "Points-Based Models for Predicting Severe Disease related to Covid-19 with the SARS-CoV-2 Omicron Variant"

### Supplementary Material

#### Supplementary Figure 1: Population Flow Chart

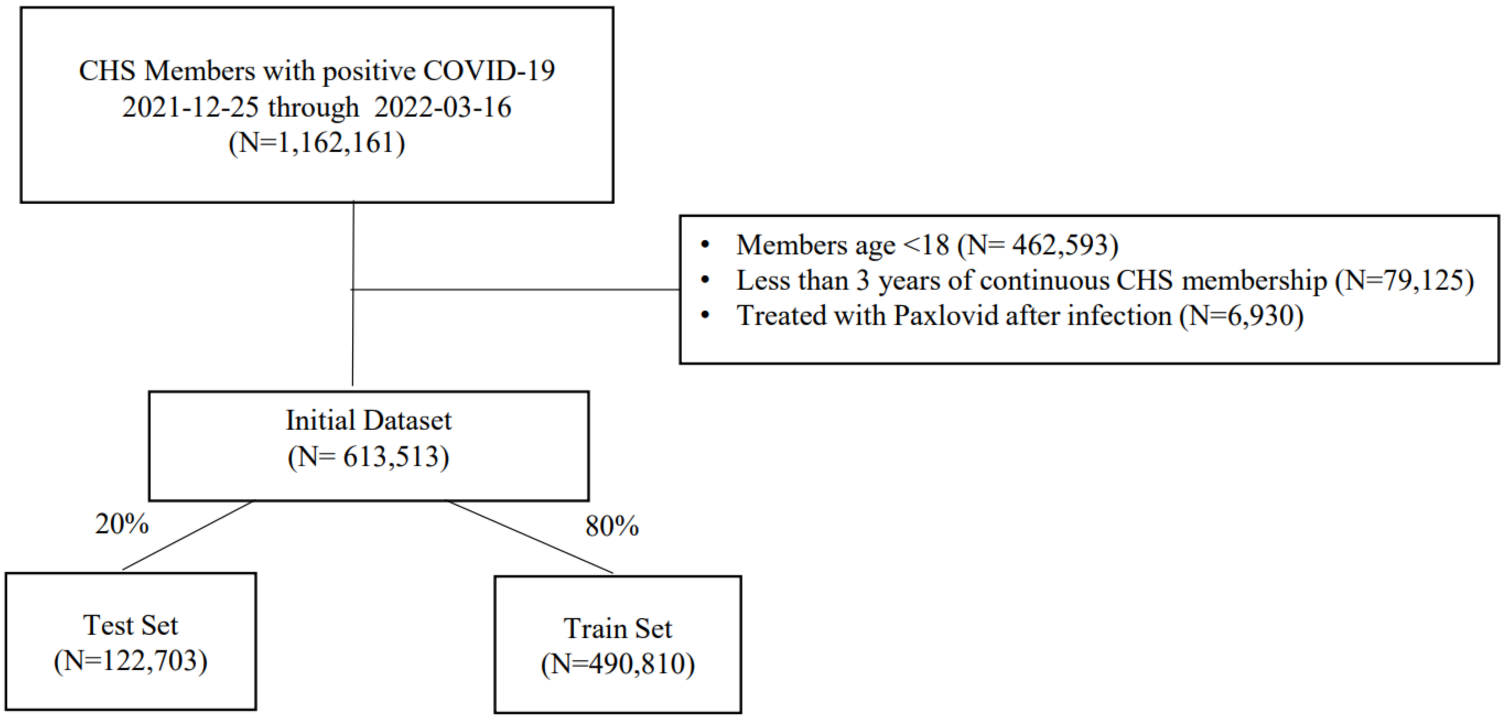

### Supplementary results

Table S1: Logistic Regression model coefficients

| Term | Coefficient | Low 95% | High 95% | P-Value | Odds Ratio | Low 95% | High 95% |
|---|---|---|---|---|---|---|---|
| (Intercept) | -7.93 | -8.53 | -7.43 | <0.001 | 0.00 | 0.00 | 0.00 |
| Age group | | | | | | | |
| 30 - 39 | 0.13 | -0.56 | 0.85 | 0.725 | 1.13 | 0.57 | 2.34 |
| 40 - 49 | 1.38 | 0.81 | 2.03 | <0.001 | 3.98 | 2.24 | 7.62 |
| 50 - 59 | 2.52 | 1.98 | 3.15 | <0.001 | 12.40 | 7.22 | 23.22 |
| 60 - 69 | 3.42 | 2.90 | 4.03 | <0.001 | 30.51 | 18.11 | 56.37 |
| 70 - 79 | 4.44 | 3.93 | 5.05 | <0.001 | 85.12 | 50.79 | 156.68 |
| 80+ | 5.45 | 4.94 | 6.06 | <0.001 | 232.50 | 139.09 | 427.22 |
| Obesity | 0.23 | 0.11 | 0.35 | <0.001 | 1.26 | 1.12 | 1.41 |
| Active cancer | 0.55 | 0.40 | 0.71 | <0.001 | 1.74 | 1.49 | 2.03 |
| Chronic Kidney Disease | 0.34 | 0.17 | 0.51 | <0.001 | 1.41 | 1.18 | 1.67 |
| Diabetes Mellitus | 0.21 | 0.04 | 0.37 | 0.012 | 1.23 | 1.05 | 1.45 |
| Heart Disease | 0.13 | -0.01 | 0.27 | 0.073 | 1.14 | 0.99 | 1.31 |
| Neurological Disease | 0.28 | 0.10 | 0.45 | 0.002 | 1.32 | 1.11 | 1.56 |
| Liver Disease | 0.27 | -0.07 | 0.59 | 0.104 | 1.32 | 0.93 | 1.81 |
| Chronic Obstruction Pulmonary Disease | 0.32 | 0.01 | 0.62 | 0.040 | 1.38 | 1.01 | 1.86 |
| Immunosuppression | 0.73 | 0.57 | 0.88 | <0.001 | 2.07 | 1.77 | 2.41 |
| Number of hospital admissions in previous 3 years | | | | | | | |
| 1 - 2 | 0.73 | 0.60 | 0.87 | <0.001 | 2.08 | 1.81 | 2.40 |
| 3 - 5 | 1.48 | 1.31 | 1.66 | <0.001 | 4.41 | 3.70 | 5.26 |
| 6+ | 2.02 | 1.79 | 2.25 | <0.001 | 7.54 | 5.99 | 9.45 |
| Months from last vaccine dose (2nd, 3rd or 4th) | | | | | | | |
| 1 - 6 | -2.13 | -2.26 | -2.00 | <0.001 | 0.12 | 0.11 | 0.14 |
| 7 - 10 | -1.58 | -1.83 | -1.35 | <0.001 | 0.21 | 0.16 | 0.26 |
| 11+ | -0.95 | -1.15 | -0.75 | <0.001 | 0.39 | 0.32 | 0.47 |
| Previous infection | -1.00 | -1.21 | -0.79 | <0.001 | 0.37 | 0.30 | 0.45 |

Table S2: Comparison of the points models in the main analysis (excluding Covid-19 therapy treated patients) with the same points models in the sensitivity analysis (including Covid-19 therapy treated patients)

| Variable | Full Points Model (Main analysis) | Simplified Points Model (Main analysis) | Full Points Model (sensitivity analysis) | Simplified Points Model (sensitivity analysis) |
|---|---|---|---|---|
| **Age group** | | | | |
| 18-29 | 21 | 7 | 21 | 7 |
| 30-39 | 25 | 8 | 23 | 8 |
| 40-49 | 31 | 10 | 31 | 10 |
| 50-59 | 39 | 13 | 38 | 13 |
| 60-69 | 45 | 15 | 44 | 15 |
| 70-79 | 51 | 17 | 50 | 17 |
| 80+ | 57 | 19 | 56 | 19 |
| **Obesity** | 2 | 1 | 1 | 1 |
| **Active cancer** | 3 | 1 | 3 | 1 |
| **Chronic Kidney Disease** | 3 | 1 | 2 | 1 |
| **Diabetes Mellitus** | 1 | 1 | 1 | 1 |
| **Heart disease** | 1 | 1 | 1 | 1 |
| **Neurological disease** | 2 | 1 | 2 | 1 |
| **Liver disease** | 3 | 1 | 2 | 1 |
| **Chronic Obstruction Pulmonary Disease** | 2 | 1 | 3 | 1 |
| **immunosuppression** | 5 | 2 | 5 | 2 |
| **Number of hospital admissions in previous 3 years** | | | | |
| 1 - 2 | 5 | 2 | 4 | 1 |
| 3 - 5 | 10 | 3 | 9 | 3 |
| 6+ | 13 | 4 | 13 | 4 |
| **Months from last vaccine dose (2nd, 3rd or 4th)** | | | | |
| 1 - 6 | -14 | -5 | -13 | -4 |
| 7 - 10 | -10 | -3 | -10 | -3 |
| 11+ | -7 | -2 | -6 | -2 |
| **Previous infection** | -7 | -2 | -7 | -2 |

Table S3 – Definitions for variables used in the study.

Variables were defined using internal CHS registries, ICD9 codes and ATC codes.

Abbreviations: MOH: Ministry of Heath; CHS: Clalit Health Services; ICD: International Classification of Disease; ATC: Anatomic therapeutic chemical;

| Variable | Values | Definitions[1] | Timing |
|---|---|---|---|
| **Outcome** | | | |
| COVID-19 related severe state (or death) | 0/1 | As defined by the hospitalizing institution per the Israeli MOH guidelines, consistent with the NIH criteria for severe illness or critical illness:<br><br>Individuals who have SpO2 <94% on room air at sea level, a ratio of arterial partial pressure of oxygen to fraction of inspired oxygen (PaO2/FiO2) <300 mm Hg, respiratory frequency >30 breaths/min, or lung infiltrates >50%.<br>Critical Illness: Individuals who have respiratory failure, septic shock, and/or multiple organ dysfunction. | |
| **Covariates** | | | |
| Cancer | 0/1 | ICD9 Code 174*<br>ICD9 Code 175*<br>ICD9 Code 233.0<br>ICD9 Code V10.3<br>ICD9 Proc Code 85.4*<br>ICD9 Code 153*<br>ICD9 Code 154*<br>ICD9 Code V10.5*<br>ICD9 Code V10.6*<br>ICD9 Code 185<br>ICD9 Code V10.46<br>ICD9 Code 162*<br>ICD9 Code V10.1*<br>ICD9 Code 188*<br>ICD9 Code V10.51<br>ICD9 Code 183*<br>ICD9 Code V10.43<br>ICD9 Code 179<br>ICD9 Code 182*<br>ICD9 Code V10.42<br>ICD9 Code 157*<br>ICD9 Code 191*<br>ICD9 Code 192*<br>ICD9 Code V10.85<br>ICD9 Code 151*<br>ICD9 Code V10.04<br>ICD9 Code 172*<br>ICD9 Code V10.82<br>ICD9 Code 201*<br>ICD9 Code 200*<br>ICD9 Code 202.4*<br>ICD9 Code 204*<br>ICD9 Code 205*<br>ICD9 Code 206*<br>ICD9 Code 207.1*<br>ICD9 Code 208.1*<br>ICD9 Code 189*<br>ICD9 Code V10.52<br>ICD9 Code 160*<br>ICD9 Code 161* | Last 5 years |

| | | | |
|---|---|---|---|
| | | ICD9 Code 164.0<br>ICD9 Code 195.0<br>ICD9 Code V10.21<br>ICD9 Code V10.22<br>ICD9 Code 180*<br>ICD9 Code V10.41<br>ICD9 Code 140*<br>ICD9 Code 141*<br>ICD9 Code 142*<br>ICD9 Code 143*<br>ICD9 Code 144*<br>ICD9 Code 145*<br>ICD9 Code 150*<br>ICD9 Code V10.03<br>ICD9 Code 155*<br>ICD9 Code 156*<br>ICD9 Code V10.07<br>ICD9 Code 170*<br>ICD9 Code V10.81<br>ICD9 Code 193<br>ICD9 Code V10.87<br>ICD9 Code 171*<br>ICD9 Code 176*<br>ICD9 Code 184*<br>ICD9 Code 186*<br>ICD9 Code 187*<br>ICD9 Code V10.4*<br>ICD9 Code 203*<br>ICD9 Code 273.3<br>ICD9 Code 152*<br>ICD9 Code 158*<br>ICD9 Code 159*<br>ICD9 Code 163*<br>ICD9 Code 164*<br>ICD9 Code 165*<br>ICD9 Code 181<br>ICD9 Code 190*<br>ICD9 Code 192.8<br>ICD9 Code 196*<br>ICD9 Code 197*<br>ICD9 Code 198*<br>ICD9 Code 199* | |
| Chronic Kidney Disease | 0/1 | ICD Proc Code 39.95<br>ICD Proc Code 54.98<br>ICD9 Code 996.81<br>ICD9 Code V42.0<br>ICD Proc Code 55.6*<br>ICD9 Code 403._1<br>ICD9 Code 404._2<br>ICD9 Code 404._3<br>ICD9 Code 585*<br>ICD9 Code 586<br>ICD9 Code 250.4*<br>ICD9 Code 274.1*<br>ICD9 Code 440.1<br>ICD9 Code 581*<br>ICD9 Code 582*<br>ICD9 Code 583*<br>ICD9 Code 587<br>ICD9 Code 588*<br>ICD9 Code 589* | Ever |
| Chronic Obstructive Pulmonary Disease | 0/1 | ICD9 Code 491*<br>ICD9 Code 492*<br>ICD9 Code 496 | Ever |
| Obesity | 0/1 | Body Mass Index (BMI) 30 or higher | Latest measurement in last 5 years not taken during pregnancy |
| Diabetes Mellitus | 0/1 | HbA1C > 6.5<br>ATC Codes A10[A,B]<br>ICD9 Code 250*<br>ICD9 Code 357.2 | For diagnosis codes, Ever |

| | | | |
|---|---|---|---|
| | | ICD9 Code 362.0* | For drugs, 4 or more dispensed in last 12 months |
| Heart Disease<br><br><br><br><br><br><br><br><br><br><br><br><br><br><br><br><br><br><br>(Including Cerebrovascular Disease) | 0/1 | ICD9 Code 410*<br>ICD9 Code 411*<br>ICD9 Code 412<br>ICD9 Code 413*<br>ICD9 Code 414*<br>ICD9 Code 429.2, 429.7*<br>ICD9 Code V45.81, V45.82<br>ICD9 Proc Code 36.0*<br>ICD9 Proc Code 36.1*<br>ICD9 Code 428*<br>ICD9 Code 398.91<br>ICD9 Code 402._1<br>ICD9 Code 404._1,<br>ICD9 Code 404._3<br>ICD9 Code 416.9<br>ICD9 Code 514<br>ICD9 Code 425*<br>ICD9 Code 416*<br><br>ICD9 Code 362.34<br>ICD9 Code 430<br>ICD9 Code 431<br>ICD9 Code 432*<br>ICD9 Code 433*<br>ICD9 Code 434*<br>ICD9 Code 435*<br>ICD9 Code 436*<br>ICD9 Code 438* | Ever |
| Immunosuppression | 0/1 | Any of:<br><br>ICD9 Code 996.81<br>ICD9 Code V42.0<br>ICD Proc Code 55.6*<br>ICD9 Code V42.7<br>ICD Proc Code 50.5*<br>ICD9 Code V42.1<br>ICD9 Code V43.2<br>ICD Proc Code 37.5<br>ICD9 Code V42.83<br>ICD Proc Code 52.8*<br>ICD9 Code V42.6<br>ICD Proc Code 33.5*<br>ICD Proc Code 33.6<br>ICD9 Code 042*<br>ICD9 Code 043*<br>ICD9 Code 044*<br>ICD9 Code 795.71<br>ICD9 Code V08<br>ICD9 Code V42.8*<br>ICD9 Proc Code 41.0*<br><br>Or at least 2 of:<br>ATC4 Code H02AB<br>ATC4 Code H02BX<br>ATC4 Code M01BA<br><br>Or at least 2 of:<br>ATC2 Code L04 | For diagnosis codes, Ever<br>For drugs, 2 or more dispensed in last 12 months |
| Neurological disease | 0/1 | ICD9 Code 290.*<br>ICD9 Code 294*<br>ICD9 Code 310.1<br>ICD9 Code 331*<br>ATC Codes N06DA02, N06DA03<br>ICD9 Code 358*<br>ICD9 Code 332.[0,1]<br>ICD9 Code 345*<br>ICD9 Code 340<br>ATC Codes L03AB07, L03AB08, L04AA07 | For diagnosis codes, Ever<br>For drugs, 4 or more dispensed in last 12 months |

| | | | |
|---|---|---|---|
| | | ICD9 Code 343*<br>ICD9 Code 333.4<br>ICD9 Code 334*<br>ICD9 Code 356*<br>ICD9 Code 138<br>ICD9 Code 335*<br>ICD9 Code 730.7*<br>ICD9 V12.02<br>ICD9 Code 228.02<br>ICD9 Code 307.23<br>ICD9 Code 330.9<br>ICD9 Code 331.3*<br>ICD9 Code 331.4<br>ICD9 Code 333*<br>ICD9 Code 334*<br>ICD9 Code 336*<br>ICD9 Code 337<br>ICD9 Code 335.1*<br>ICD9 Code 359.0<br>ICD9 Code 359.21<br>ICD9 Code 357.0<br>ICD9 Code 237.7*<br>ICD9 Code 742.8[1,2] | |
| Liver Disease | 0/1 | ICD9 Code 070.22<br>ICD9 Code 070.23<br>ICD9 Code 070.32<br>ICD9 Code 070.33<br>ICD9 Code 070.44<br>ICD9 Code 070.54<br>ICD9 Code V02.61<br>ICD9 Code V02.62<br>ICD9 Code 571*<br>ICD9 Code 275.1<br>ICD9 Code 277.4<br>ICD9 Code 452<br>ICD9 Code 453.0<br>ICD9 Code 571.8<br>ICD9 Code 571.9<br>ICD9 Code 572* | Ever |
| hospital admissions | Categorical Variable 0, 1-2, 3-6, 6+ (months) | Count of any inpatient hospitalization excluding: Same-day hospitalizations, dialysis treatments, normal delivery, psychiatric hospitalizations and long-term care (nursing) hospitalizations | In the last 3 years |
| Previous infection | 0/1 | A previous PCR or Antigen confirmed SARS-CoV-2 infection. Occurring at least 90 days prior to the current infection. | Ever |

Abbreviations: MOH, Ministry of Heath; ICD, International Classification of Disease; ATC, Anatomic therapeutic chemical;

[1]Additional confirmation of the diagnostic codes was done by checking the matching of the free text within the diagnosis description field.
